# Development and evaluation of wrist- and thigh-worn accelerometer algorithms using self-training machine learning models for classification of activity type and posture: towards device placement-agnostic methods in the ProPASS consortium

**DOI:** 10.1101/2025.06.26.25330373

**Authors:** Matthew N. Ahmadi, Nicholas Koemel, Raaj Biswas, Andreas Holtermann, Annemarie Koster, Andrew Atkin, Richard Pulsford, Borja del Pozo Cruz, Vegar Rangul, John Mitchell, Joanne Blodgett, Pasan Hettiarachchi, Magnus Svartengren, Malcom Granat, Bronwyn Clark, Mark Hamer, Emmanuel Stamatakis

**Affiliations:** University of Sydney; The University of Sydney; National Research Centre for the Working Environment; Maastricht University; The University of East Anglica; University of Exeter; University of Southern Denmark; Norwegian University of Science and Technology; University College London; Uppsala University; University of Salford; University of Queensland; UCL

## Abstract

**Background:** Wearable accelerometers are widely used in health research, but differing placements (e.g. wrist vs. thigh) hinder harmonising activity classification across studies. Prior studies report 1.5- to 2.0-fold differences in physical activity level by wear location, hampering data comparability, and compromising the potential for pooling data to develop consortia and carrying out Meta- and Individual Participant Data analysis. Although supervised machine learning is increasingly used in wearables research, its reliance on extensive labelled data limits its use in free-living datasets. Semi-supervised learning offers an efficient alternative by using laboratory collected labelled data to iteratively self-train models on unlabelled free-living data. Using a self-training approach, the aim of this study was to train and evaluate algorithms for wrist- and thigh-worn devices to facilitate harmonisation of posture and activity type classification between placements.

**Methods:** A total of 146 participants aged 30-75 years completed either structured laboratory-based activity trials or one of two independent free-living assessments while wearing Axivity AX3 accelerometers on the wrist and thigh. For each placement, a supervised Random Forest classifier was initially trained using a labelled laboratory dataset (n=40) to classify sitting, standing, walking, running, stair climbing, and cycling - and then re-trained using self-training on free-living data (n=53, independent to the laboratory study sample). The final models were validated using another hold-out free-living independent dataset (n=53) with ground-truth activity labels obtained via direct video observation. Overall model comparison and performance was assessed using accuracy, kappa statistic, and F1 scores. Individual activity class comparison and performance was evaluated using equivalence testing, confusion matrices, and coefficient of variation between the wrist and thigh estimates.

**Results:** During a total of 43,800 minutes, of which 19,080 minutes were in the hold-out dataset, both self-trained models achieved high overall classification accuracy: 91.8% (SD = 6.8%) for the wrist and 95.1% (SD = 5.4%) for the thigh. The overall F1 score was 88.2 (SD = 9.6%) for the wrist classifier and 90.1 (SD = 9.3%) for the thigh classifier. Equivalence testing demonstrated that both classifiers produced activity duration estimates statistically equivalent to ground-truth for all activity types except stair climbing. Confusion matrices for the wrist demonstrated very good to excellent (88% - 97%) classification accuracy for sitting, walking, running, and cycling, and good accuracy for standing and stair climbing (71%–78%). For the thigh, classification performance was very good to excellent (83% - 98%) across sitting, standing, walking, running, and cycling, with good accuracy for stair climbing (75%). The coefficient of variation values ranged from 0.022 for running to 0.140 for standing.

**Conclusion:** These findings highlight the potential of self-training models to support harmonisation of wearable accelerometer data collected using different wear placements in ProPASS and other consortia. Self-training models reduce reliance on extensive labelled data and demonstrated high activity type classification accuracy for both wrist- and thigh-worn accelerometers, with a high degree of agreement and equivalence with ground-truth data across almost all activity types.

## Introduction

Wearable devices, primarily featuring accelerometer sensors, are a common assessment method to quantify physical activity in observational and intervention studies ^1,2^. To date, wearables have been used to characterise limited aspects of physical activity behaviour; primarily restricted to intensity-based metrics or total physical activity amount or volume. Contemporary studies are utilising wearables to provide more comprehensive outputs that characterise other aspects of physical activity, such as activity type and posture (sit vs stand)^3–6^ which can be used to provide greater details about potential health benefits^7^ and to fully capture 24-hour behaviours^6^. While the capacity of wearables to measure a broader range of dimensions of physical activity advancing, the longstanding issue of harmonising the output of accelerometry data from different body placements remains unresolved^8,9^. Notably, a recent systematic review reported over the past decade, most studies have used wearables placed on either the wrist or the thigh^10^. However, physical activity measures derived from these two placements are currently considered incompatible, which hinders the integration of data in modern evidence synthesis resources—such as Individual Participant Data (IPDs)^11^ that require pooling of different population cohorts, intervention studies, and consortia like ProPASS^6,12–14^.

In the absence of a systematically developed harmonisation methodology, previous attempts to pool data from multiple wear placements have resulted in disparate activity duration estimates. For example, studies have reported a 1.5- to 2.0-fold difference in physical behaviour estimates between placements, leading to incongruent estimates of activity prevalence^15^ and associations with health outcomes^16^. This was due, in part, to previous processing techniques that were designed for a specific wear placement site. For example, the magnitude of acceleration is a reliable indicator of whole-body movement when the device is worn near the body’s centre of mass, such as the hip or thigh. However, the magnitude of acceleration does not accurately capture whole body movement activity when applied to wrist-worn devices, where it primarily reflects localized arm motion^17^. As the World Health Organization moves towards integrating wearables into the next iteration of global surveillance^18^ and future guidelines development^19^, it is crucial to develop methodologies that can be used to harmonise physical activity estimates across different wear placements. To address these challenges, collaborative initiatives have emerged. The ProPASS consortium^12^ was established in 2017 as a thigh-worn accelerometry-focused initiative to specifically serve the next generation of physical activity guidelines through large scale IPDs, spearheading the transition from self-reported evidence to wearables-based data^20^. As of 2023, ProPASS has expanded to include cohorts with wrist-worn accelerometry data.

Machine learning can enhance the utility of wearables by leveraging rich sensor data to harmonize physical activity measures across placements. Prior studies using supervised learning models reported less than 10% differences between placements but rely on extensive labelled data (ground-truth) collected in controlled laboratories where ground-truth data is easily accessible. However, when applied to free-living conditions, this leads to a 25–40% reduction in accuracy due to the unstructured nature of daily activities^21,22^. Training models on large volumes of free-living data with ground-truth is challenging due to logistical constraints in cost and time, such that few models of this nature exist^23–25^. Semi-supervised learning offers a promising alternative by leveraging a small amount of labelled data to develop a base classifier, which is then iteratively retrained on large volumes of unlabelled free-living data.

This process of using the model’s own predictions to refine itself, known as self-training, could—if validated across diverse conditions—help bridge the gap between controlled laboratory environments and the complexities of real-world settings, ultimately enhancing the generalizability and utility of wearable sensor data.

Previous applications of self-training for natural language processing, object detection, and image classification have shown improvements in classification accuracy compared to traditional supervised learning classifiers^26–29^. Importantly, recent studies have demonstrated self-training techniques can be applied to supervised learning classifiers^30–35^, such as Random Forest classifiers. If found to be effective for activity recognition with wearables, this approach could improve the generalizability of machine learning models which would significantly facilitate harmonisation of accelerometer data collected using different wear placements.

Using a semi-supervised self-training approach, the aim of this study was to train and evaluate algorithms for wrist- and thigh-worn devices to facilitate harmonisation of posture and broad activity type recognition between placements. Additionally, we compared the performance of these new models to traditional supervised classification models.

## Methods

### Participants

A total of 146 participants between the ages of 30 to 75 years participated in the three independent arms of the study. Participants were recruited through university e-mailing lists, social media, and word of mouth.

Interested individuals were provided with an information sheet explaining the study and eligibility criteria. Participants were excluded from the study if they met any of the following exclusion criteria: 1) had neurological or musculoskeletal impairments limiting daily activities; 2) had health conditions or used medications affecting heart rate response to physical activity; 3) had diagnosed respiratory disease; 4) implanted cardiac medical devices; 5) had stable angina pectoris; 6) indicated any contraindications after completion of the Physical Activity Readiness Questionnaire (PAR-Q)^36^. This study was approved by the Sydney Local Health District (ETH02815, ETH00138). Before participation, written consent was obtained from each participant.

### Data collection

Participants were allocated to one of three study arms.

#### Arm 1: Base-model - laboratory data collection

This dataset was used to train base models on labelled ground-truth data and comprised of 40 participants (61.3 +/- 12.7 y; 20 female) who completed a 90-minute activity session. During the sessions, participants wore an Axivity AX3 (Newcastle upon Tyne, UK) accelerometer on their dominant wrist and on the anterior aspect of their right thigh, midway between the greater trochanter (hip) and the lateral epicondyle of the femur. Each participant completed the following activity trials: 1) Lying supine; 2) sitting and reading or writing; 3) washing dishes at a kitchen sink; 4) carrying items approximately 10-15 meters from the kitchen to a table; 5) treadmill walking at a self-selected comfortable pace; 6) energetic activities (such as playing soccer or basketball); 7) cycling; 8) stair climbing; 9) running or fast walking.

Each activity trial lasted for 5 minutes and activity start and stop times were recorded using a Polar wrist-watch that was initialised on the same computer used to initialise the Axivity device to ensure timestamp synchronisation. Accelerometer data was annotated with activity type using the activity start and stop times. By design, the activity trials incorporated physical activity typically performed in day-to-day life by adults, and included both locomotor and intermittent/lifestyle activities.

#### Arm 2: Self-training - free-living data collection

This dataset was used to retrain the base models on unlabelled (e.g. no ground-truth) free-living data and comprised 53 participants (55.4 +/- 13.1 y; 35 female) who completed a 6-hour free-living data collection session during which they engaged in their usual daily activities in the community while wearing an Axivity AX3 on their dominant wrist and right thigh. The participants also wore a body-worn video recorder (Losoform Z02; Guangzhou, China) to attain direct observation ground-truth data of the time and duration of activities performed. Video files were imported into the Noldus Observer XT 15 software (Noldus Information Technology, Wageningen, the Netherlands) for continuous direct observation coding. A previously established direct observation scheme^37^ was implemented in which the participant’s movement behaviour was coded as one of six activity classes using activity types from the physical activity compendium. These posture and broad activity classes were: 1) sitting; 2) standing; 3) walking; 4) stair climbing; 5) running/high energetic activities; and 6) cycling. The computerised direct observation system generated a vector of date–time stamps corresponding to start and finish of each movement event, which were used to calculate event duration and assign the activity type to the corresponding time segments of the accelerometer data. Interobserver reliability was assessed by dual coding 24 randomly selected videos across 5 video coders. The intraclass correlation coefficient was 0.902 (95% confidence interval = 0.872 – 0.932).

#### Arm 3: Hold-out - free-living data collection

This dataset was used to evaluate the performance of the models and comprised another 53 participants (57.4 +/- 13.2 y; 31 females) who completed a 6-hour free-living data collection session wearing the same monitors as Arm 1 and Arm 2 participants on their dominant wrist and right thigh. Participants also wore the same body-worn camera as described above with the same direct observation coding scheme applied to the recordings.

### Model development and evaluation: training, self-training, and testing

Devices were calibrated and non-wear periods were detected using standard procedures^38,39^. Time- and frequency-domain features from the x, y, and z axis were extracted from 10s non-overlapping windows. This window length was chosen to provide sufficient time resolution to detect physical activity movement patterns. A total of 75 features previously shown to be beneficial in activity recognition were extracted^40–43^.

A Random Forest classifier was trained using labelled data from Arm 1 (laboratory) to classify posture and activities into one of six classes: 1) sitting; 2) standing; 3) walking; 4) stair climbing; 5) running/high energetic activities; and 6) cycling. This classifier served as a base-model that was then applied to Arm 2 (self-training on free-living data).

In the Arm 2 dataset, we applied self-training techniques where the ground-truth labels were removed so the dataset became an “unlabelled dataset”. Self-training involves two phases: 1) The base classifier, trained on labelled data, is used to predict labels for the unlabelled dataset; 2) The predicted labels that have high prediction confidence, referred to as ‘pseudo-labels,’ are then used to retrain the model, thereby enhancing its ability to generalize to new, unobserved (e.g. free-living) data^44^. If the prediction confidence was ≥80%, the predicted activity class was used to retrain the classifier. The self-training process continued iteratively until the improvement in prediction confidence across iterations was <2%, indicating convergence.

Lastly, the sequence of activity probabilities by the Random Forest classifier was smoothed over time with a Hidden Markov Model (HMM) to produce the final sequence of activity predictions. The HMM classifier was trained using ground-truth data obtained from Arm 2 participants. HMM can improve the accuracy of predictions by modelling the likelihood of transitions between activities over time. For instance, it accounts for the improbability of transitioning directly from sitting to cycling without an intermediate activity such as walking. Additionally, the HMM places greater weight on activity predictions with higher confidence, thereby reducing the influence of less certain predictions during the smoothing process. Models were trained using the ‘randomForest’, “HMM”, and ‘DMwR2’ packages in R.

The wrist and thigh retrained Random Forest classifiers were tested on Arm 3 participants (hold-out – free-living data collection) using the video direct observation as the ground-truth labels. This hold-out dataset was entirely separate from the Arm 1 (laboratory training) and Arm 2 (self-training) datasets, ensuring that the evaluation was not influenced by any overlap in the data used for training. By using ground-truth labels derived from direct observation, the evaluation provided a reliable benchmark for assessing the classifiers’ ability to generalize to real-world scenarios. The hold-out dataset was essential for assessing the true predictive accuracy of the models and their capacity to classify activities across different free-living contexts.

The use of a separate hold-out dataset aligns with best practices in machine learning, as it ensures the evaluation results are reflective of the models’ performance on new data. This rigorous testing approach is critical when developing methodologies intended for deployment in free-living environments, to assess the robustness and scalability of the models for real-world applications.

#### Model evaluation and statistics

To assess overall performance of the wrist and thigh models, we calculated accuracy, kappa statistic, and F1-scores (to account for class imbalance in Arm 3).

Equivalence testing^45–47^ was conducted to determine whether the activity class predictions for the wrist model and thigh model were statistically equivalent to ground-truth measures. Unlike traditional null hypothesis testing, which aims to show that there is no significant difference, equivalence testing is used to demonstrate that two methods produce results that are equivalent within a pre-specified equivalence margin—defined by lower and upper bounds—indicating that any differences within these bounds are clinically insignificant. In our study, we set both the lower and upper bounds at a Cohen’s d of 0.3, representing a small-to-medium effect size. This threshold corresponds to an approximate 80% overlap between the distributions^48^ of physical activity estimates from a classifier when compared to ground-truth. By establishing equivalence between wrist and thigh outputs to ground truth, this approach can help confirm if sensor placement does or does not introduce meaningful discrepancies in activity classification, thereby supporting the harmonisation of wearable data for pooling large-scale population studies and behavioural trials.

To complement the equivalence testing and quantify the magnitude and consistency of agreement between the wrist and thigh, we further calculated the absolute standard deviation (AbsSD) and coefficient of variation (CV). For each participant, we calculated the SD between wrist and thigh activity and posture durations. The AbsSD is the average of the SD across all participants providing a measure of absolute disagreement (in minutes). The CV provides the relative variability (ratio of SD / mean) across all participants and reflects proportional disagreement between placements. Low AbsSD and CV values demonstrate minimal practical variability between activity and posture estimates across both placements^49^.

To determine how each classifier performed in detecting individual activity types against ground-truth, we generated heatmap confusion matrices. These matrices are used to identify misclassification patterns and serve as the basis for calculating class-level sensitivity, specificity, and F1-scores. Performance was further evaluated using Bland-Altman plots to assess agreement between each model (wrist and thigh) and ground-truth data in estimating time spent in each activity class. This analysis allowed us to identify any systematic bias, quantify the limits of agreement, and examine the distribution of differences at the participant level, thereby providing insights into the consistency and reliability of the classifiers.

To compare the performance of the self-trained models with traditional supervised classification models reliant on only labelled data, a supervised Random Forest classifier (for the wrist and thigh) was trained using the ground-truth labelled data from Arm 2 and evaluated using Arm 3.

## Results

Participant characteristics for the three arms are presented in **Table 1**. Arm 1, used to train the base classifier on laboratory data, included 40 participants (20 female) with a mean age of 61.3 (12.7) years, 21 of whom were overweight or obese. Arm 2, used for self-training of the base classifier on unlabelled free-living data, comprised 53 participants (31 female) with a mean age of 57.4 (13.2) years, of which 31 were overweight or obese. The hold-out dataset (Arm 3), used to evaluate wrist and thigh model performance, consisted of another 53 participants (35 female) with a mean age of 55.4 (13.1) years, with 31 classified as overweight or obese. The self-trained and traditional supervised Random Forest wrist and thigh classifiers were built with 500 trees, and the number of features randomly sampled at each split was optimized at 8. The self-trained wrist and thigh classifiers are available as part of the ProPASS open-source repository at https://github.com/Ergo-Tools/ActiPASS.

**Table 1:** Participant characteristics for the 3 datasets.

|  | **Arm 1** | **Arm 2** | **Arm 3** | **Overall** |
| --- | --- | --- | --- | --- |
| **N** | 40 | 53 | 53 | 146 |
| **Age** | 61.3 (12.7) | 57.4 (13.2) | 55.4 (13.1) | 57.6 (13.2) |
| **Females** | 20 | 31 | 35 | 86 |
| **Height, cm** |  |  |  |  |
| Males | 173.8 (7.7) | 175.7 (5.3) | 176.9 (3.9) | 175.5 (5.8) |
| Females | 162.6 (6.6) | 162.7 (7.1) | 162.9 (7.6) | 162.8 (7.1) |
| **Weight, Kgs** |  |  |  |  |
| Males | 84.3 (16.6) | 83.8 (11.8) | 83.3 (9.4) | 83.7 (12.7) |
| Females | 61.4 (12.2) | 68.1 (13.4) | 68.6 (17.1) | 66.8 (15.1) |
| **Body Mass Index (%)** |  |  |  |  |
| Normal | 19 (47.5%) | 22 (41.5%) | 22 (41.5%) | 63 (43.2) |
| Overweight | 15 (37.5%) | 22 (41.5%) | 19 (35.9%) | 59 (40.4) |
| Obese | 6 (15%) | 9 (17%) | 12 (22.6%) | 24 (16.4) |

Values represent mean (SD) unless noted otherwise
Arm 1 was used to train the base model on labelled data
Arm 2 was used to re-train the models on free-living unlabelled data
Arm 3 was used to evaluate the models on free-living data with ground-truth video direct observation

**Table 2** displays the mean ± SD of time spent in each activity and posture per participant in the hold-out dataset (Arm 3). On average, based on ground truth data, participants spent 194 (± 54) minutes sitting, 106 (± 50) minutes standing, 39 (± 24) minutes walking, 4 (±1) minutes stair climbing, 3 (± 1) minute running/high energetic activities, and 12 (± 27) minutes cycling. Average AbsSD (CV values in parentheses) values were 6.1 (0.78) for sitting, 11.0 (.140) for standing, 2.3 (.063) for walking, 0.2 (.041) for stair climbing, 0.4 (.022) for running/high energetic activities, and 1.6 (.083) for cycling.

**Table 2:**
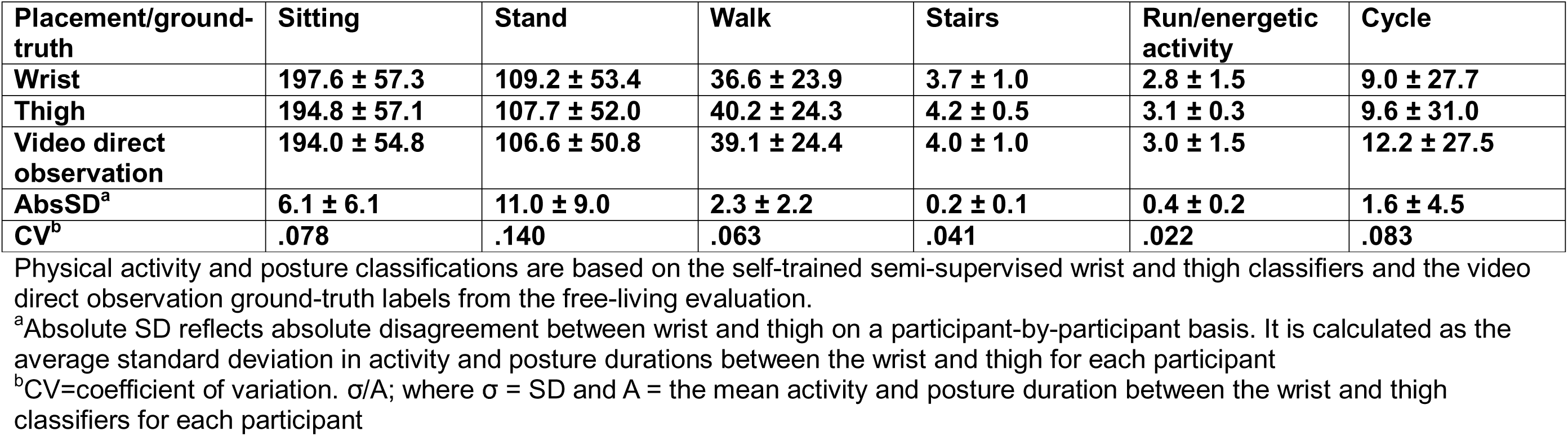
Physical activity and posture duration (minutes) per participant in free-living evaluation (Arm 3)

| **Placement/ground-truth** | **Sitting** | **Stand** | **Walk** | **Stairs** | **Run/energetic activity** | **Cycle** |
| --- | --- | --- | --- | --- | --- | --- |
| **Wrist** | **197.6 ± 57.3** | **109.2 ± 53.4** | **36.6 ± 23.9** | **3.7 ± 1.0** | **2.8 ± 1.5** | **9.0 ± 27.7** |
| **Thigh** | **194.8 ± 57.1** | **107.7 ± 52.0** | **40.2 ± 24.3** | **4.2 ± 0.5** | **3.1 ± 0.3** | **9.6 ± 31.0** |
| **Video direct observation** | **194.0 ± 54.8** | **106.6 ± 50.8** | **39.1 ± 24.4** | **4.0 ± 1.0** | **3.0 ± 1.5** | **12.2 ± 27.5** |
| **AbsSD<sup>a</sup>** | **6.1 ± 6.1** | **11.0 ± 9.0** | **2.3 ± 2.2** | **0.2 ± 0.1** | **0.4 ± 0.2** | **1.6 ± 4.5** |
| **CV<sup>b</sup>** | **.078** | **.140** | **.063** | **.041** | **.022** | **.083** |

Physical activity and posture classifications are based on the self-trained semi-supervised wrist and thigh classifiers and the video direct observation ground-truth labels from the free-living evaluation.
<sup>a</sup>Absolute SD reflects absolute disagreement between wrist and thigh on a participant-by-participant basis. It is calculated as the average standard deviation in activity and posture durations between the wrist and thigh for each participant
<sup>b</sup>CV=coefficient of variation. $\sigma/A$ ; where $\sigma$ = SD and A = the mean activity and posture duration between the wrist and thigh classifiers for each participant

### Overall performance

Participant level overall accuracy for the self-trained Random Forest wrist classifier was 91.8% (SD= 6.8%) with a Kappa statistic of 81.4 (SD= 12.1) and an overall F1 score of 88.2 (SD= 9.6). In comparison, the self-trained Random Forest thigh classifier had an overall accuracy of 95.1% (5.4%), with a Kappa statistic and F1 score of 90.2 (10.6) and 90.1 (9.3), respectively. Individual activity class sensitivity, specificity, and F1 score are shown in **Supplemental Table 1**. For sitting, walking, stair climbing, and running, class-level sensitivity, specificity, and F-scores for the wrist and thigh were within 5% of each other with the thigh consistently showing closer agreement to ground-truth for posture.

### Equivalence testing

Equivalence results for time spent in different activity classes are displayed in **Figure 1**. The wrist and thigh self-trained Random Forest classifier demonstrated equivalence with ground-truth video direct observation for all six activity or posture classes, except for stair climbing. Although the confidence interval for stair climbing crossed zero for both placements, it also crossed the upper bound, indicating that while the prediction was not statistically different from the ground-truth, it was not statistically equivalent either. For the wrist classifier, sitting and standing were overestimated by 3.5 (90% CI = −26.3, 33.40) and 2.6 (−12.1, 17.3) minutes respectively. Walking had a mean underestimation of 1.3 minutes, with a 90% CI ranging from −10 to 12 minutes. For the thigh classifier, the corresponding values for sitting, standing, and walking were 0.8 (−24.9, 26.5) minutes, 2.6 (−12.1, 17.3) minutes, and 1.1 (−10.2, 12.4) minutes.

**Figure 1:**
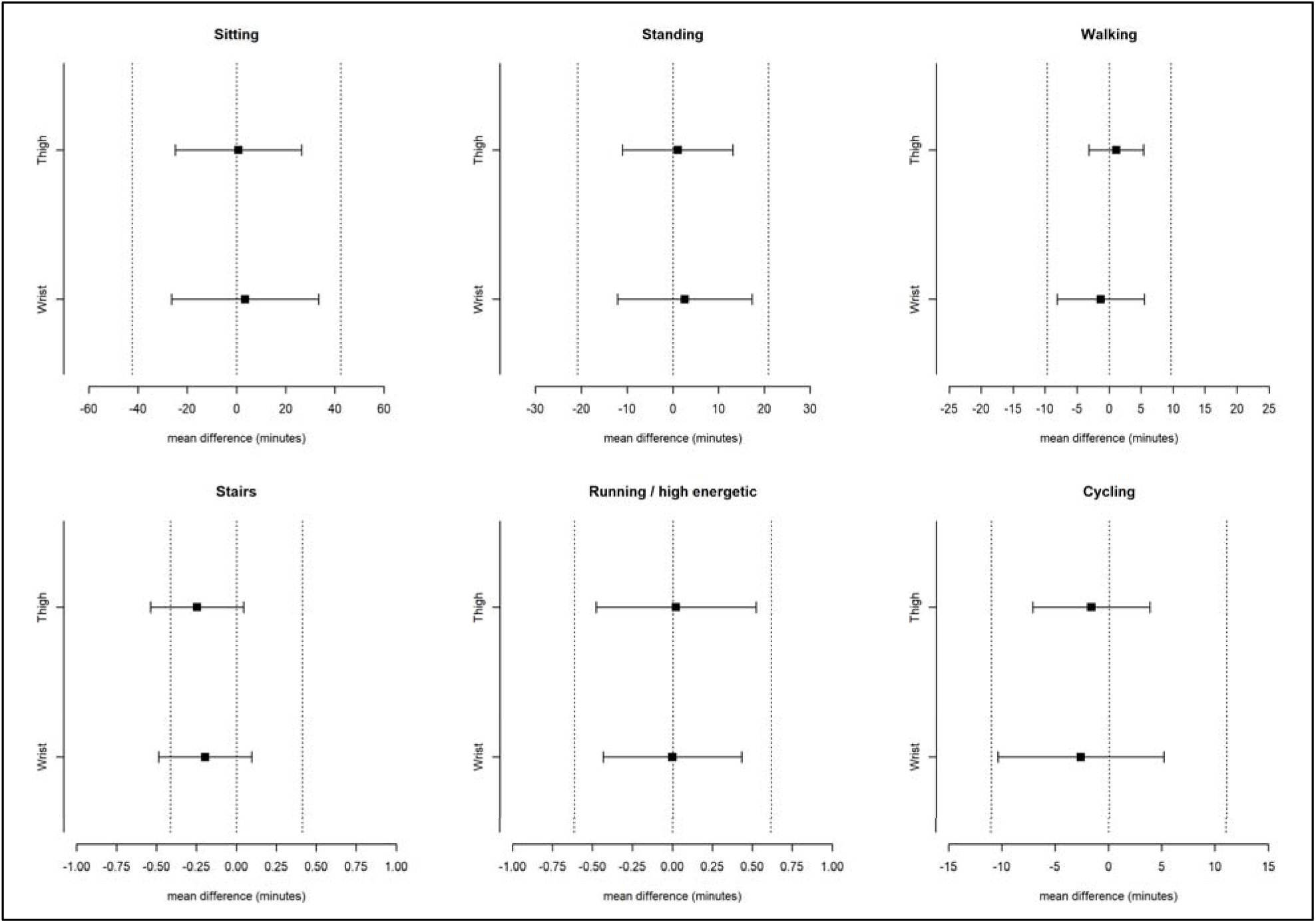
Equivalence of wrist and thigh classifiers to ground-truth data Equivalence testing of thigh and wrist to ground-truth. Positive values indicate overestimation and negative values indicate underestimation

### Confusion matrices

Figures 2-3 shows the heatmap confusion matrix for the self-trained Random Forest classifiers. For the wrist classifier (Figure 2), recognition of sitting, walking, running/high energetic, and cycling were very good to excellent ranging from 88% to 97%. Recognition of standing and stair climbing was good ranging from 71% to 77%. The majority of misclassification for wrist captured standing was when it registered as sitting during periods of standing still, and the majority of stair climbing misclassification occurred when it was registered as walking during periods of combined stair climbing and walking within a given 10 second window period. For the thigh classifier (Figure 3), recognition of sitting, standing, walking, and running/high energetic activities was excellent and ranged from 91% to 98%.

**Figure 2:**
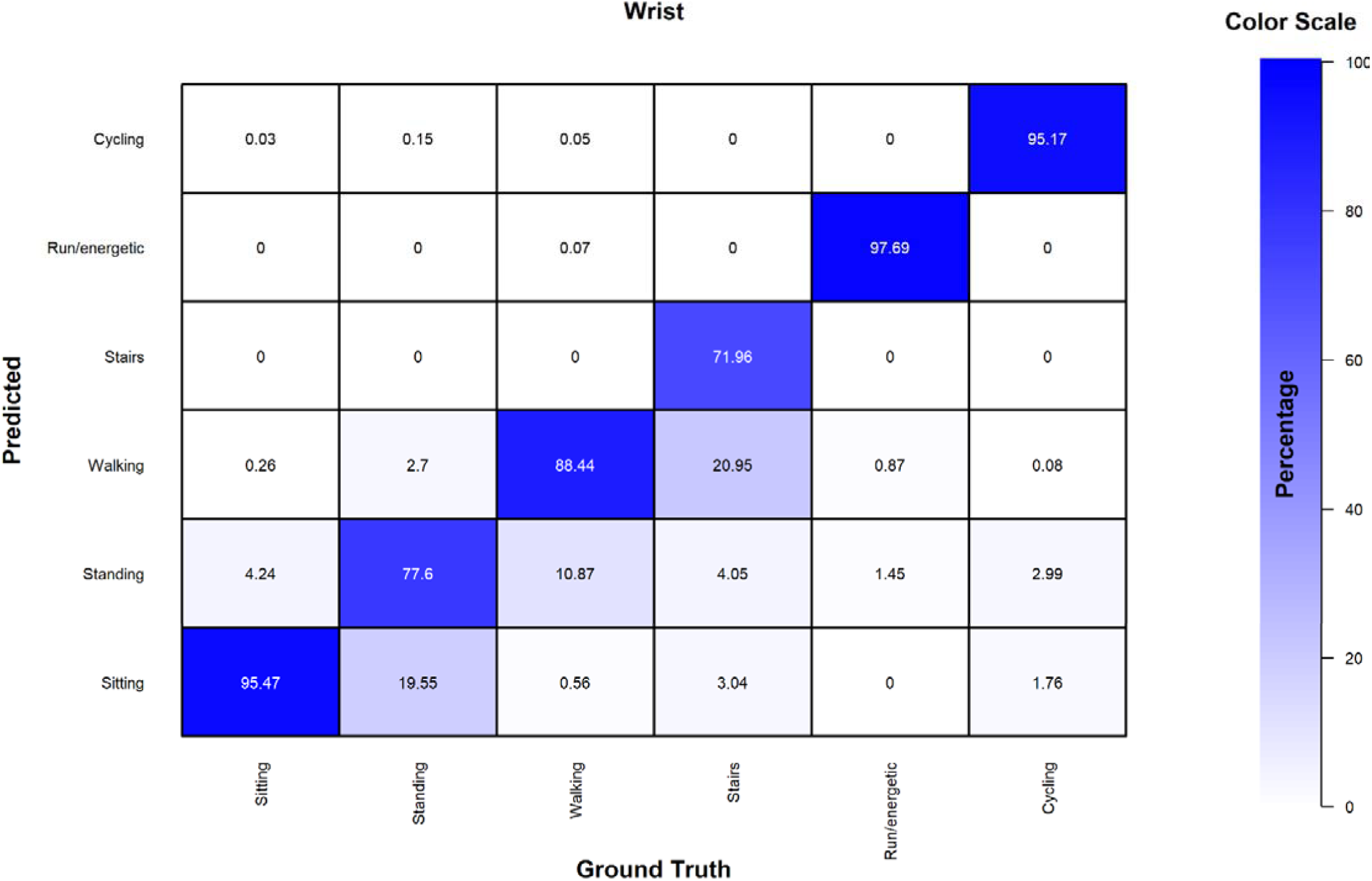
Heatmap confusion matrix for the wrist classifier

**Figure 3:**
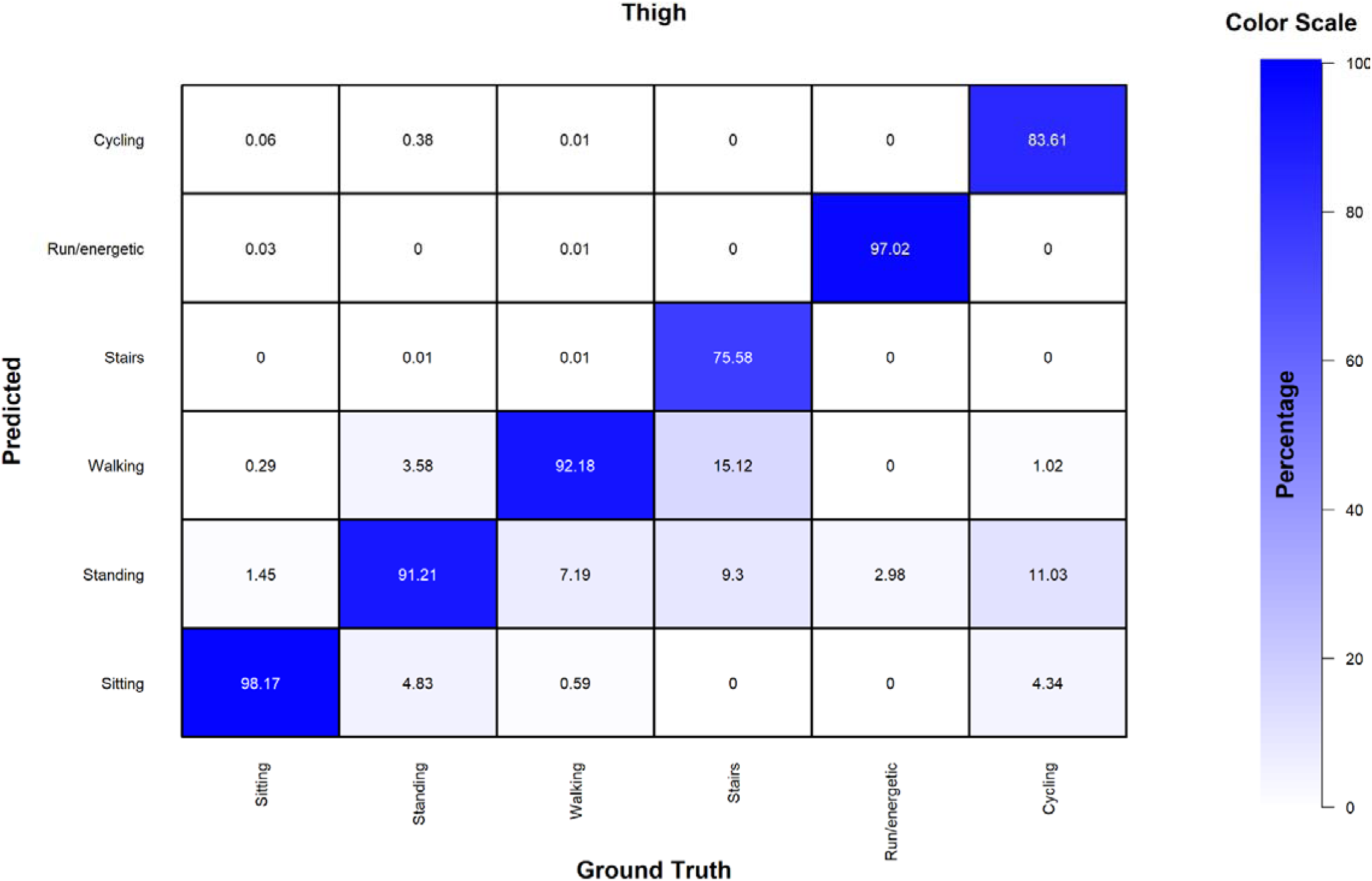
Heatmap confusion matrix for the thigh classifier

Recognition of thigh stair climbing was moderate to good (76%) and cycling was good to very good ranging from 75% to 83%. The majority of misclassification for thigh captured stair climbing was when it was registered as walking and the majority of misclassification for cycling occurred as standing – during periods when the participant had intermittently stopped pedalling, to free-wheel.

### Bland-Altman analysis

The Bland-Altman plots (Figure 4) indicate the self-trained Random Forest classifiers for the wrist and thigh generally had good agreement for time spent in different activities and postures with ground-truth video direct observation. Both classifiers demonstrated minimal mean bias (e.g. <5 minutes) and no systematic bias across each of the activities and postures. For both placements, walking and running/high energetic activities demonstrated the strongest alignment with ground-truth showing narrow limits of agreement. Sitting and standing exhibited more variability for the wrist with limits of agreement between −26 minutes to 31 minutes. For the same two activity classes, the thigh classifier had limits of agreement between −13 minutes to 16 minutes. To aid interpretation, **Supplemental Table 2** presents the absolute differences as relative (percentage) differences, offering a complementary perspective on the degree of deviation from ground-truth across activities and postures.

**Figure 4:**
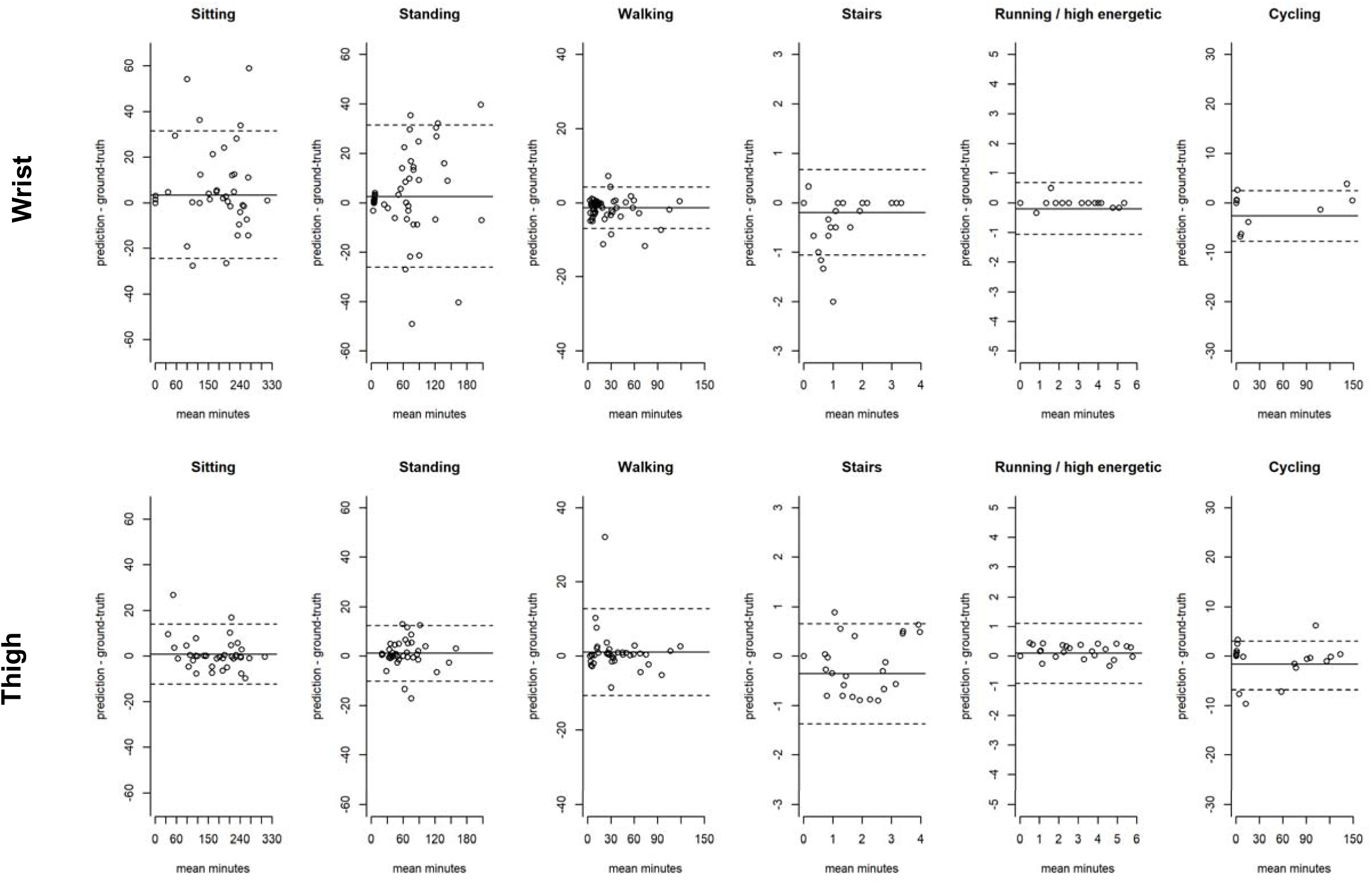
Bland-Altman plots for time spent in different activity classes

### Comparison to traditional supervised Random Forest classifier

**Supplemental** Figures 1–2 display the confusion matrices for the traditional supervised Random Forest classifiers for both wrist and thigh placements, and **Supplemental Tables 3–4** show the corresponding class-level sensitivity, specificity, and F-scores. The confusion matrices revealed similar classification and misclassification patterns compared to the self-trained classifiers. For the wrist classifier, classification accuracy ranged from 74% for stair climbing to 98% for running. For the thigh classifier, accuracy ranged from 81% for stair climbing to 98% for running and sitting. For individual activity classes, sensitivity, specificity, and F-scores differed by no more than 5% from those of the self-trained classifiers for each respective placement.

## Discussion

The self-trained activity classifiers developed and evaluated in this study demonstrates considerable potential to harmonise activity type and posture data from wearables worn on the wrist and thigh. It also highlights the value of leveraging self-trained semi-supervised learning techniques in combination with free-living ground truth data, which collectively may open new possibilities for developing classifications suitable for pooling of wearables data in consortia such as ProPASS^12^ and other consolidated resources of observational or clinical trials data.

Advancements in harmonisation methodologies could enable scalable and efficient activity classifier development approaches, thereby supporting large-scale analyses that fully exploit the rich information captured by wearable devices.

We evaluated activity classifiers for both wrist and thigh placements, the most common wear locations in epidemiological studies over the past decade^10^. Given the prevalence of these placements, developing harmonised classification models is essential to ensure that activity and posture estimates are consistent and reliable across studies using different protocols. Without such harmonisation, discrepancies in device placement introduce bias, compromising the validity of pooled analyses and hindering evidence synthesis. Our work addresses this challenge by demonstrating that harmonised classifiers can provide comparable activity and posture estimates, supporting robust cross-study comparisons and facilitating meta-analyses, and ultimately advancing the application of physical activity data to improve public health insights and interventions.

Harmonising physical activity and posture measurements across wrist- and thigh-worn devices will enable the integration of diverse international datasets, resulting in at least two key benefits. First, it will improve the precision of physical activity and posture assessments in population surveillance and interventions studies, enhancing our understanding of health effects and policy-relevant prevalence alike^8^. Second, it may support more detailed and robust observational analyses of physical activity and posture patterns and their associations with mortality and non-communicable diseases. The ability to pool international, population-level wearable data,-regardless of differing wear protocols, can advance the field toward developing wearables informed physical activity guidelines and policies aimed at improving physical activity and posture behaviours ^1,2^. Beyond population health, standardising harmonisation procedures and wearable data processing protocols will support more precise assessments, enabling targeted interventions and precision medicine strategies.

In addition, our findings offer methodological flexibility to new studies to choose an accelerometer placement that is most cost-effective, more likely to produce highest wear adherence among participants, and ultimately yield the highest quality data. For this purpose, it is still important to consider that each placement has unique strengths, e.g. thigh placements are more accurate for posture allocation while wrist-worn placements have slightly better feasibility^10^. Ultimately, these advances will improve how population-level physical behaviours and health outcomes are monitored and researched.

Unlike traditional supervised models that require large amounts of labelled data—an often time-intensive and resource-heavy process— our approach effectively incorporates both labelled and unlabelled data, significantly reducing the dependency on labour-intensive annotation efforts^50,51^. By applying self-training on data collected in real-world environments, the models were able to adapt and learn movement patterns that are more reflective of free-living conditions. Our application of self-training techniques showed classifiers for different wear placements provided similar activity estimates to free-living ground-truth data. Previous studies have reported that physical activity estimates can differ by 1.5- to 2.0-fold depending on wear placement when relying solely on acceleration magnitude^16^. Additionally, supervised models trained exclusively on free-living data have been shown to yield differences greater than 10% across placements in detecting common activities such as walking or running^40^.

While self-training techniques have been previously applied to Random Forest classifiers^29,52^, our study is the first to specifically adapt these methodologies for physical activity and posture classification. We leverage flexible data harmonisation techniques^53^, which allow for the integration of data collected using diverse protocols (such as different device placements) by applying systematically developed harmonisation methodologies^54^. Unlike stringent harmonisation, which requires identical measures and procedures across datasets, flexible harmonisation focuses on making variables-such as physical activity and posture-collected using different protocols (e.g., device placement) inferentially equivalent^55^, enabling synthesis across a broad range of activities, postures, and study designs. In our study, this was demonstrated through equivalence testing, showing that harmonised data from different wear placements could be meaningfully compared to free-living ground-truth data. The strong consistency between activity estimates from self-trained wrist and thigh classifiers support pooling data from cohorts using different wear placements.

Such standardised methodology, including those used in Network harmonization^56^, will help ensure that observed differences in activity type are attributable to genuine behavioural variation rather than sensor placement. Although we did not observe statistical equivalence for stair climbing, we found minor differences in absolute duration and low coefficient of variation (0.041 equivalent to 4.1%). The negligible size of these differences indicate that they are unlikely to be of practical relevance in population-level studies, yet identifies an area requiring further exploration.

Future work should expand on our activity type and posture harmonisation by developing methods to harmonise wrist and thigh device assessments for physical activity intensity, sleep duration/quality, and step count. While the coefficient of variation for standing between the thigh and wrist models was 0.14 (14%), indicating moderate relative variability in their estimates, application of ComBat^57^ (combining batches) and CovBat^58^ (covariance batch effects) techniques can be used. ComBat can harmonise mean and variance differences in acceleration features across the wrist and thigh, reducing placement-specific biases in posture classification. CovBat extends this by harmonising on multivariate features (e.g. frequency components) to harmonise their distributions between placements. An important next step is to create scalable, open-source, and flexible harmonisation pipelines, such as those based on federated learning frameworks^59,60^, which would enable consortia like ProPASS to collaboratively train adaptable machine learning models on internationally diverse datasets without the need to share raw data, thereby ensuring participant privacy and data security.

### Strengths and limitations

Our study had several strengths. We demonstrated the feasibility and utility of self-training in harmonising wrist and thigh models and allowing them to adapt to new, unobserved movement patterns and perform efficiently in real-world environments. Additionally, our use of a video direct observation scheme provided continuous ground-truth data for activity types. This approach enabled us to capture very short and intermittent activities, as well as to identify multiple activities occurring in rapid succession, thereby offering valuable insights into misclassification patterns. There are also several limitations that warrant consideration. First, signal features were extracted using a fixed sliding window which may lead to multiple activity classes occurring within a single window, and an underestimation of time spent in some activity classes. Future studies should explore the use of alternative windowing techniques or point detection. Second, our models used features extracted from a single accelerometer sensor. Utilisation of sensor fusion to combine data from multiple sensors capturing both movement and physiological information, may improve activity type accuracy and warrants further exploration in future research.

## Conclusion

By leveraging self-training semi-supervised learning techniques, free-living ground truth data, and flexible harmonisation methodologies, we demonstrated a high degree of agreement between wrist and thigh accelerometer placements for quantifying physical activity and posture types. Our findings support the feasibility and scientific rigour of pooling wrist and thigh wearables data when such classification methods are employed. Semi-supervised learning reduces reliance on labour-intensive labelled datasets, aligning with broader trends in wearables research that prioritise scalable, real-world data utilisation to develop machine learning models. These improvements in harmonisation and model development methodologies will enable large population consortia such as ProPASS to pool data, as well as prospective meta-analyses and IPDs to fully leverage wearables data to strengthen physical activity and health research.

## Data Availability

All data produced are available online at https://github.com/Ergo-Tools/ActiPASS

