## Supplemental Document for "Development and evaluation of wrist- and thigh-worn accelerometer algorithms using self-training machine learning models for classification of activity type and posture: towards device placement-agnostic methods in the ProPASS consortium"

**Supplemental Table 1:** Individual activity class sensitivity, specificity, and F1 score

Wrist classifier

| **Activity or posture** | **Sensitivity** | **Specificity** | **F-score** |
| --- | --- | --- | --- |
| Sitting | 95.5 | 89.3 | 94.0 |
| Standing | 77.6 | 94.6 | 79.0 |
| Walking | 88.4 | 99.0 | 91.1 |
| Stairs | 72.0 | 100 | 83.7 |
| Running | 98.0 | 99.9 | 97.8 |
| Cycling | 95.2 | 99.9 | 97.6 |

Thigh classifier

| **Activity or posture** | **Sensitivity** | **Specificity** | **F-score** |
| --- | --- | --- | --- |
| Sitting | 98.2 | 96.7 | 98.0 |
| Standing | 91.2 | 97.1 | 90.4 |
| Walking | 92.2 | 98.8 | 92.1 |
| Stairs | 75.6 | 99.9 | 85.0 |
| Running | 97.0 | 99.9 | 96.0 |
| Cycling | 83.6 | 99.9 | 89.7 |

**Supplemental Table 2:** Bland-Altman results displaying relative percentage differences for mean bias and 95% Limits of Agreement

| **Activity** | **Wrist** | **Thigh** |
| --- | --- | --- |
| Sitting | 1.9 (-13.7, 17.7) | 0.4 (-7.2, 8.1) |
| Standing | 3.0 (-29.5, 35.4) | 1.7 (-16.6, 20.0) |
| Walking | -4.0 (-20.9, 12.9) | 3.0 (-25.2, 31.3) |
| Stair climbing | -8.5 (-46.3, 29.3) | -10.1 (-49.4, 27.1) |
| Running | -4.7 (-18.5, 17.8) | -4.9 (-21.4, 18.2) |
| Cycling | -3.1 (-5.9, 5.3) | -.3.6 (-6.2, 6.4) |

Values represent % difference from mean of ground-truth + predicted time in each activity and posture class. A positive value denotes overestimation; a negative value denotes underestimation.

**Supplemental Figure 1:** Confusion matrix for the wrist classifier trained on free-living labelled data for the traditional supervised Random Forest classifiers.

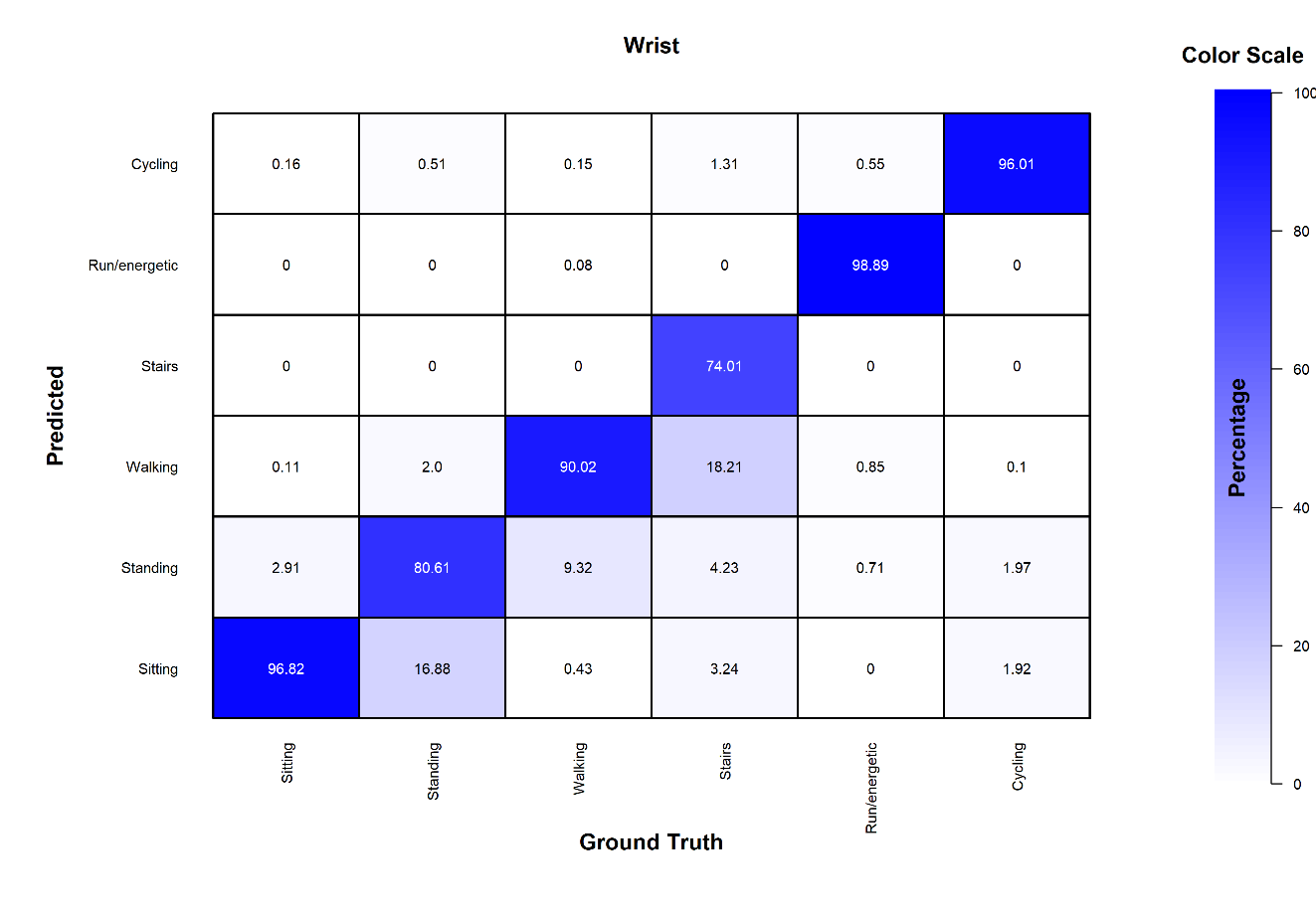

**Supplemental Table 3**

Class level sensitivity, specificity, and F-score for the traditional supervised Random Forest classifiers

| **Activity** | **Sensitivity** | **Specificity** | **F-score** |
| --- | --- | --- | --- |
| Sitting | 96.8 | 90.8 | 95.2 |
| Standing | 80.6 | 95.9 | 82.6 |
| Walking | 90.0 | 99.3 | 92.8 |
| Stairs | 74.0 | 100 | 84.6 |
| Running | 98.9 | 99.9 | 98.0 |
| Cycling | 96.0 | 99.8 | 95.0 |

**Supplemental Figure 2:** Confusion matrix for the wrist classifier trained on free-living labelled data

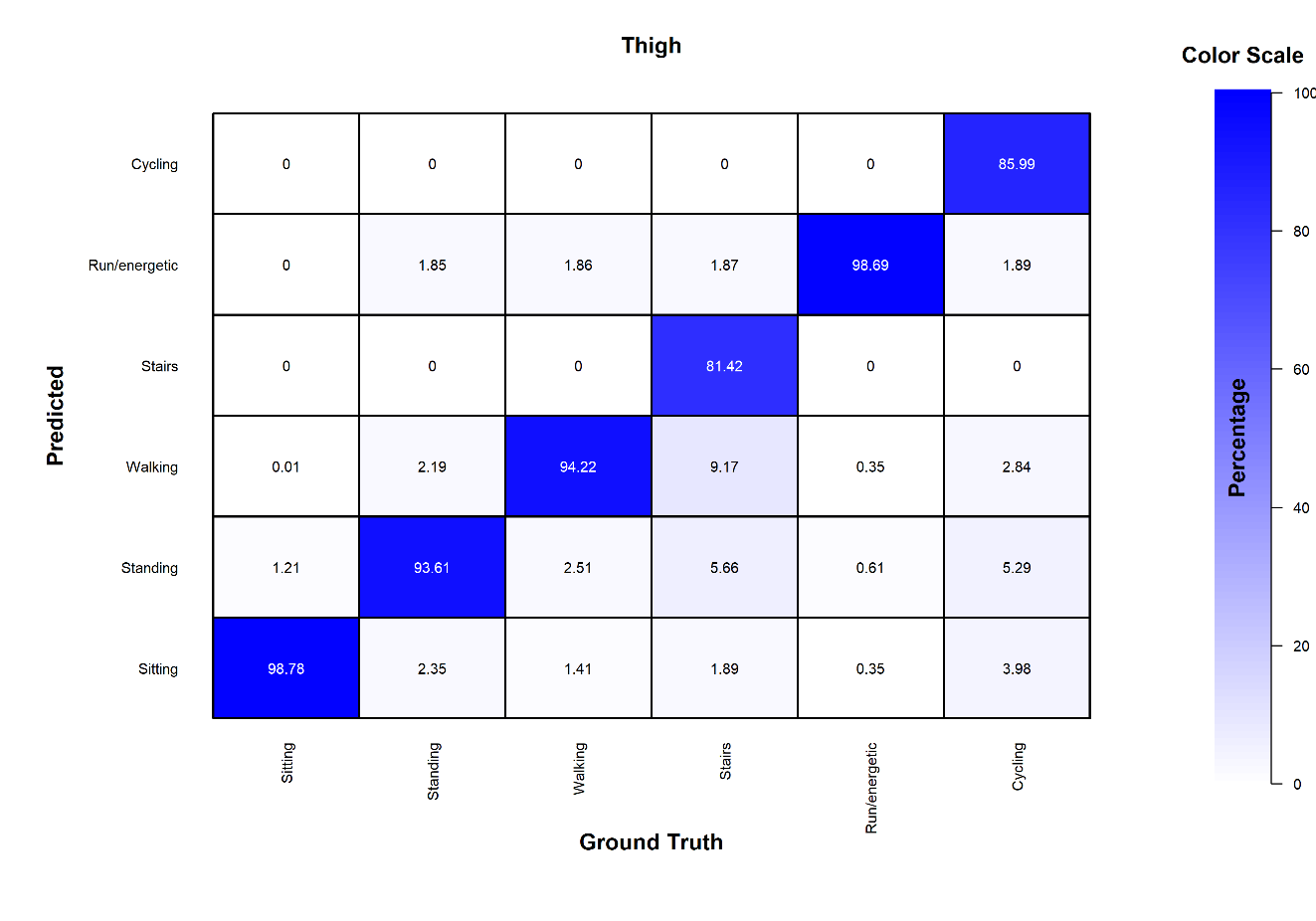

**Supplemental Table 4**

Class level sensitivity, specificity, and F-score

| **Activity** | **Sensitivity** | **Specificity** | **F-score** |
| --- | --- | --- | --- |
| Sitting | 98.8 | 97.9 | 98.6 |
| Standing | 93.6 | 98.3 | 93.9 |
| Walking | 94.2 | 99.3 | 94.8 |
| Stairs | 81.4 | 100 | 89.6 |
| Running | 98.7 | 99.2 | 58.6 |
| Cycling | 86.0 | 100 | 92.5 |
